# A disposable female urinal bottle (the EasyWee ^tm pending^) improves patient experience for immobilised females with lower limb fractures

**DOI:** 10.1101/2024.05.06.24306419

**Authors:** Siân Thomas, Sophy Booth, Peter Ellis, Savneet Lochab, Mark D Lyttle, James Pegrum

**Author notes:** **Corresponding author:** Siân Thomas, Great Western Hospital, Emergency Department, Marlborough Rd, Swindon SN3 6BB. **Contributors:** ST designed the study, is fully responsible for the conduct of the study, had access to the data, analysed the data, is responsible for drafting this manuscript and controlled the decision to publish. SB, PE, SL, JP and ST led the recruitment of participants and management of data in their respective departments. ML supervised the design of the data collection tool, provided advice on study design and contributed to manuscript writing. SB, PE, SL and JP have contributed to the final manuscript writing. All authors have read and approved the final version of the manuscript. **Funding:** No funding was received for this project. **Competing interests:** None declared. **Patient and Public Involvement:** Patients and/or public were not involved in the design, conduct, reporting or dissemination of this project. **Patient consent for publication:** Not required. **Ethics approval:** This project was undertaken as a service improvement project. Waiver of ethical approval for the first two cycles was confirmed by Dr. Donna Noonan, Head of Service, Research & Innovation, Great Western Hospitals NHS Foundation Trust. The multi-site, third cycle element of the project was approved by the Health Research Authority and Health and Care Research Wales (NHS) on 22nd March 2022 (IRAS project number 311131) with NHS Research Ethics Committee (22/PR/0191) (London-Harrow) approval. Participants gave informed consent to use the urinal and to provide anonymised feedback. **Data availability statement:** Data are available on reasonable request. Requests to the corresponding author – Siân Thomas. De-identified participant data can be made available on request. **Intellectual Property:** Intellectual property is owned by Great Western Foundation Hospitals NHS Trust. Design protection is pending for this urinal. The name “EasyWee” has trade mark protection pending.

## Abstract

Female patients with lower limb fractures often experience pain and loss of dignity when manoeuvred onto a bedpan. Poor bladder management, including urinary catheterisation for convenience, can lead to longer hospital stays and eventual loss of independence. Disposable pulp male urinal bottles have been modified into a shape that accommodates the female perineum but they have not been used consistently, the design has varied depending on the fabricator and no formal evidence supported their use.

Using the Model for Improvement process with sequential Plan, Do, Study, Act (PDSA) cycles we formalised the design and spread the use of the urinal to other hospitals and an ambulance service. This project was inspired by a patient advocate of the “female” urinal.

A local feasibility study focussed on female patients with hip fracture in an emergency department and was followed by a second cycle in both emergency and trauma departments. The final cycle was a multi-institutional study^*^. The final study cohort was immobilised female patients (n=103) and healthcare professionals supporting them (n= 118). Utility was assessed by feedback on the advantages of the urinal. Healthcare professionals were also asked about impact on their working practices. Acceptability was addressed by asking patients and healthcare professional participants whether they would recommend the urinal.

Most patients and healthcare professionals in this trial would recommend the urinal to another patient. Patients felt they suffered less pain from movement and that it was a more dignified way to micturate. Healthcare professionals felt the advantages were potential to reduce the need for urinary catheters, for lifting and log rolling patients.

A disposable urinal that accommodates female anatomy and supplies the same advantages as the male urinal bottle (ability to micturate without the need for movement) would have value in many clinical settings.

- **What is already known on this topic** – female *patients with lower limb fractures suffer pain and indignity when being moved onto bed pans to micturate, men can use a male urinal bottle without being moved*.
- **What this study adds** – *a female-specific urinal bottle is acceptable to both patients and healthcare staff, more dignified and less painful for patients and less work for healthcare staff*.
- **How this study might affect research, practice or policy** – *a female-specific urinal bottle may reduce the use of urinary catheters for immobilised females with reduction in catheter-associated infections and loss of independence. It anticipated that it will reduce lifting by healthcare staff and reduce the number of staff required to assist patients*.

## Introduction

### Problem description

There is inequity in the management of urinary care of immobilised patients without male anatomy, usually, but not exclusively female. Male patients who are immobile are offered commercially available pulp urinal bottles that can be used with minimal aid and little movement (Schluter *et al*. 2017). Female patients with lower limb fractures often suffer both pain and indignity during bladder management.

Hip fractures are the most common orthopaedic inpatient admission with the current UK incidence estimated at 76,000 per annum (National Hip Fracture Database Annual Report 2023) and around 75% of those sustaining hip fractures are female (Lisk and Yeong 2014). Fractures are painful at the time of injury and any time the patient moves or is moved (Pellino 1994) for example onto a bedpan despite optimal analgesia such as fascia iliaca block (Callear and Shah 2016). NICE recommends a multi-disciplinary hip fracture programme which includes management of pain for patients with a neck of femur fracture (NICE CG124). We therefore concentrated initially on using the urinal for female neck of femur (NOF) patients

Common practice for bladder management in female patients involves the use of a bed pan (Rodriguez 2016). Alternatively, an indwelling or intermittent urinary catheter is inserted with associated risk of infection, delirium and long-term impact on urinary continence and independence (Farrington *et al*. 2016), (Thomas *et al*. 2021). For patients who cannot tolerate either, or soil themselves, incontinence pads are used with risk of associated dermatitis and urinary tract infections (Payne 2015). Reluctance to use the bedpan can lead to avoiding fluid intake to prevent bladder filling (Gattinger *et al*. 2013), with resultant preoperative dehydration and post operative kidney injury (Ellis *et al*. 2018). The use of bed pans also increases patient dependency (Cohen 2009), and patients are often left waiting for assistance to void (Mitchell 2023), (Hughes 2008).

This project evaluates a disposable pulp female urinal that can be used by an immobilised patient when lying or sitting and is in line with National Clinical Guidelines that promote minimising pain and post operative delirium associated with hip fractures (NICE CG124/ NICE CG103).

### Available knowledge

There is limited literature advocating use of female urinals in hospital settings (Saint and Lipsky 1999) (Rodriguez and Adriel 2016) and few equivalent products designed for female anatomy. A small study of palliative patients concluded that female urinals should be available and offered to female patients in hospital (Farrington et al 2016). An abstract from a conference described a study of use of non-disposable urinals for female orthopaedic patients. Their results showed that the majority of patients found the female urinal more comfortable than a bedpan and would recommend use of the female urinal (Rodriguez and Adriel 2016). A group has published a paper describing a methodology to scan the female perineum to optimise the design of the opening of a female urination device. The limitation of this paper is that is used a small number of young Chinese women who may not be representative of the wider population (Wang et al 2015). There is a vast array of commercially available female urinals, some of which are disposable, most of which are designed for use whilst standing, some can be used sitting. At the time of the study there was one commercially available, pulp, female urinal, which is significantly more expensive than its male counterpart and used in a manner similar to that of the bedpan.

### Rationale

This urinal is a simple modification of the disposable, pulp, round urinal bottle that accommodates female anatomy for immobilised patients, with the perceived benefits of minimising pain, maintaining patient dignity and autonomy, and reducing staffing needs. This can be used with the patient lying or sitting and requires minimal or no movement of the patient. The intervention was advocated by a patient in the first instance and was therefore assumed to have a reasonable likelihood of success.

### Specific Aims

The Institute for Healthcare Improvement’s Model for Improvement process with sequential Plan, Do, Study, Act (PDSA) cycles *(Langley et al 2009)* was used to evaluate the utility and acceptability of the urinal to patients and healthcare professionals in emergency departments (ED) and trauma and orthopaedic departments (T&O) and in different hospitals (Table 1). The questions addressed were: is the modified male pulp urinal bottle useful for patients without male anatomy; is it acceptable to that cohort of patients and healthcare professionals supporting them. Our aim was to have developed an acceptable product that could be presented for commercialisation by the end of 2023.

**Table 1:**
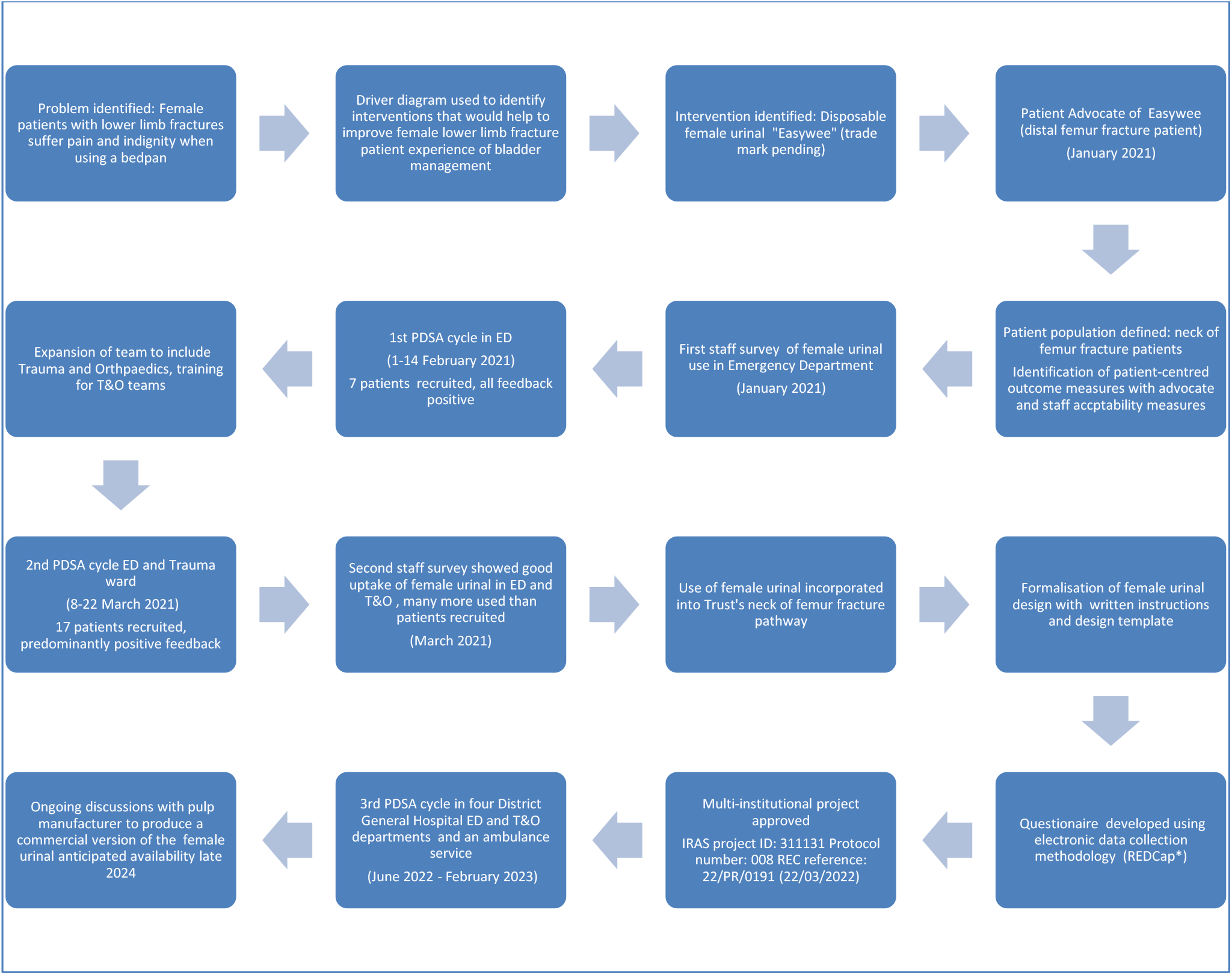
Time line of the sequential PDSA cycles used and the associated development of the female urinal project.

## Methods

### Context

The first patient advocate was sufficiently invested in the intervention to have taught her subsequent care givers in the Trauma Department how to make the urinal. A patient advocate is invaluable but to implement the use of a new urinal staff must also act as advocates. This project required healthcare staff to use and make a non-commercial product to assist patients toileting. Although many staff working in emergency departments are resourceful and practical not all staff are comfortable with the prospect of fashioning a female urinal with a pair of scissors! In some cases, it was possible to pre-prepare a box of female urinals but physical space to store extra boxes was in some cases problematic. The advantages for staff included less lifting and reduced numbers of staff to assist with toileting. A small number of staff who had found that the intervention successful through personal experience shared their knowledge with others on an ad hoc basis. The purpose of this study was to formalise and expand this knowledge base and provide evidence to support the use of this female urinal.

### Intervention

The urinal is fashioned by cutting around the neck of the pulp urinal bottle to create a wider mouth with a posterior section that can be slid under the perineum (Figure 1). The design consensus was established after the second PDSA cycle. The urinal used in the last cycle were either pre-prepared or cut for use on demand by individual healthcare workers. Guidance on how to cut the product was provided through leaflets and a ‘How-To’ video. Attempts to minimise variation in design consisted of reviewing design in sites during the project and sharing images of an “optimal” design. The urinal was primarily made by nursing staff, health care staff and emergency department assistants but were also made by paramedics and doctors.

**Figure 1:**
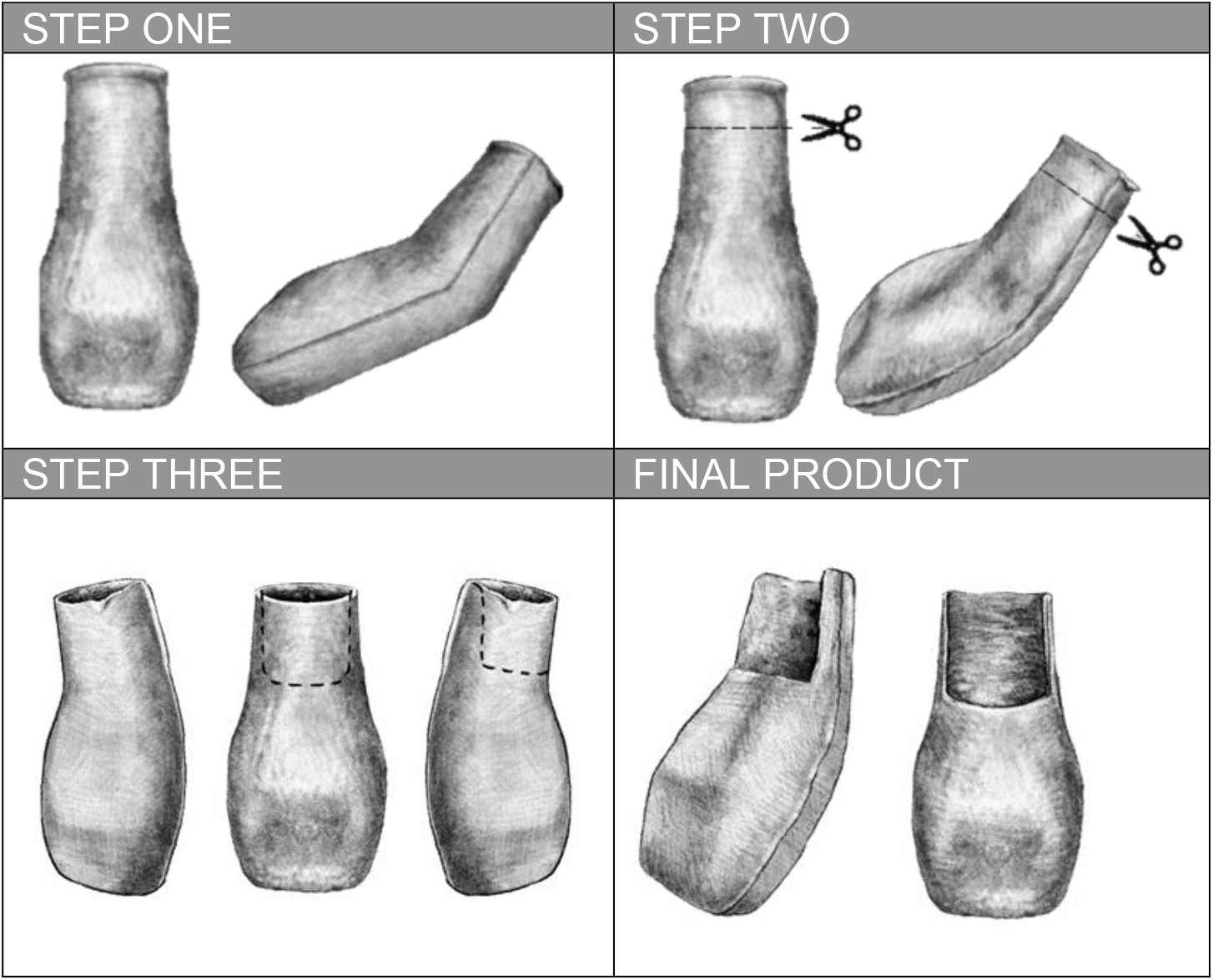
Visual representation of how to make the urinal

The initial team leading the first cycle of this project were an Emergency Department Assistant, Paramedic and Registrar from the ED with advice from the patient advocate. As the project progressed the team was expanded to include the trauma nurse lead, orthopaedic Senior House Officer and Orthogeriatric Consultant at the same hospital. The multi-institutional study had leads at each institution and included ED and Orthopaedic consultants and a Research Nurse and Ambulance Service Research Lead.

The urinal was used by an ambulance service supporting the emergency department of a site, and in emergency departments and/or trauma and orthopaedic wards of four district general hospitals based in the South West region of England.

### Intervention

The urinal was offered to patients with female anatomy who were over the age of 18, had capacity to verbally consent, were not in acute urinary retention, were immobilised (primarily those who had sustained lower limb fractures such as neck of femur fractures), were unable to use a male urine collection bottle, needed to micturate and were admitted to the study sites. All healthcare professionals providing a urinal to a patient were invited to participate.

### Measures

The primary outcome measures were utility and acceptability to patients and healthcare professionals. Utility was assessed by asking patients closed questions about the advantages of the urinal. Healthcare professionals were also asked closed questions about their perception of the utility of the urinal for patients and the impact on their working practices. Acceptability was addressed by asking patients and healthcare professionals whether they would recommend the urinal. Free text boxes were provided. Healthcare professionals were asked to provide limited demographic information about the patient. Patient position and BMI were collected by healthcare professionals as these variables may affect utility or acceptability (Table 2).

**Table 2:**
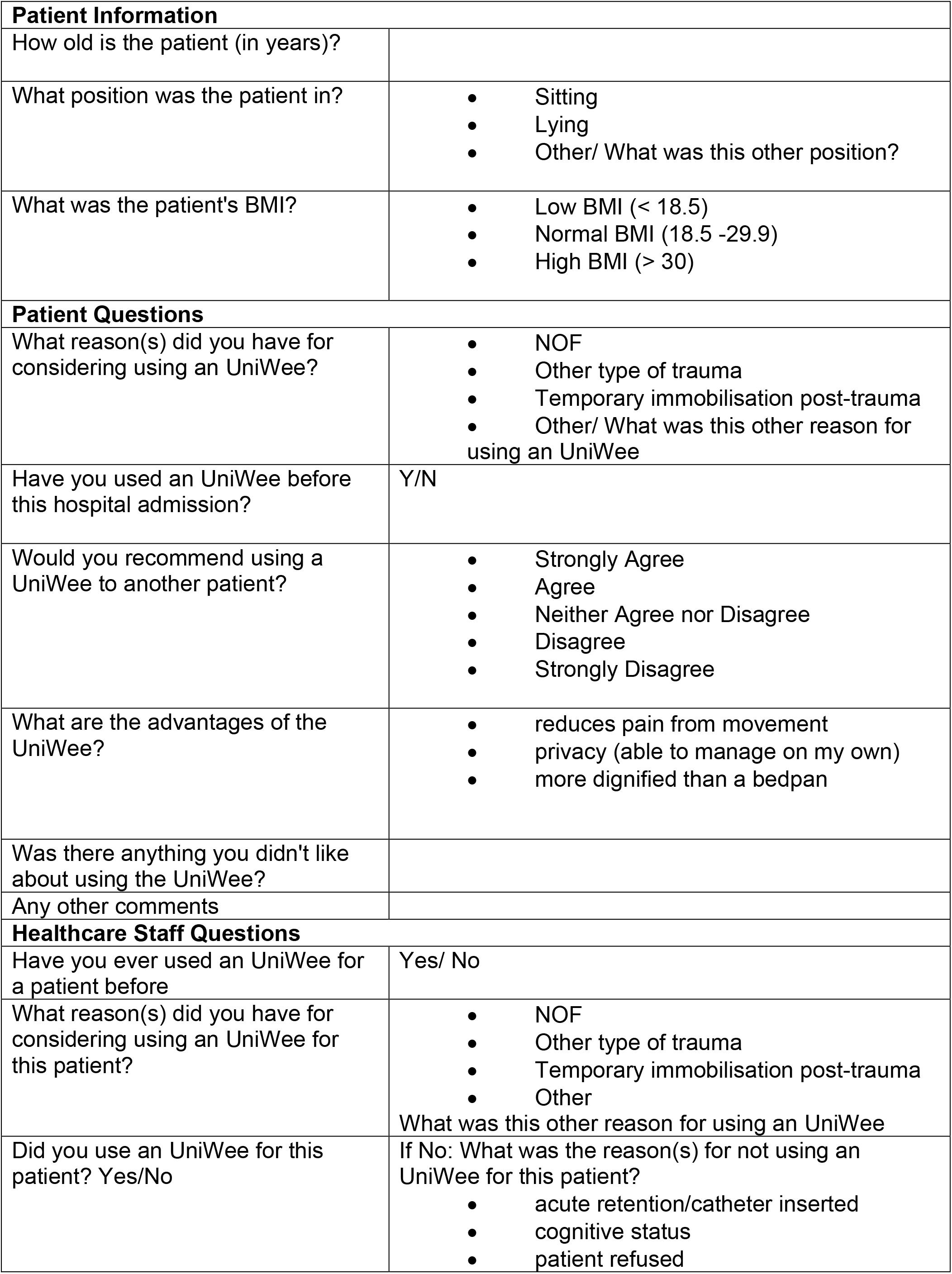
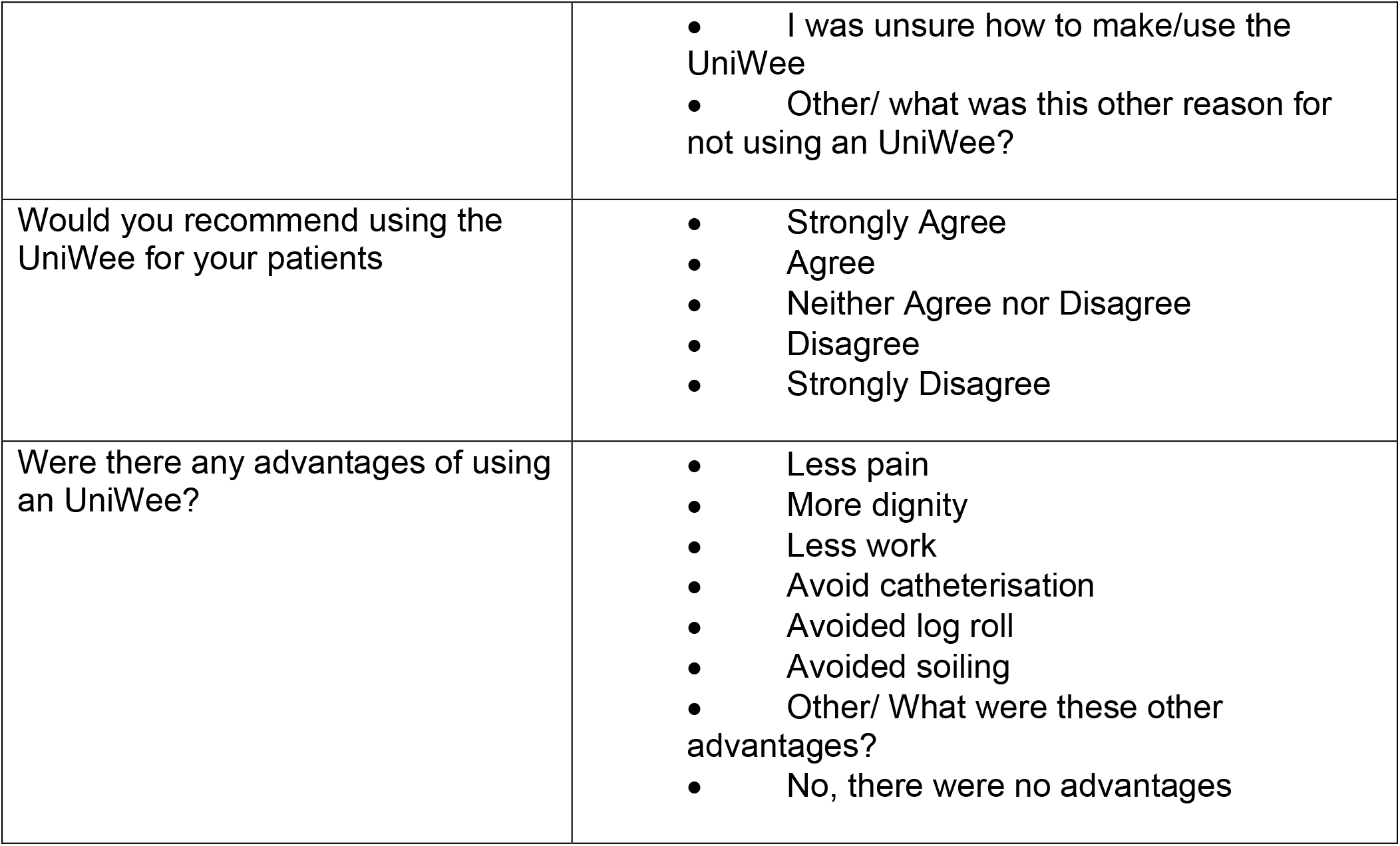
Measures used for assessing the outcomes of using the UniWee.

|  |  |
| --- | --- |
| **Patient Information** |  |
| How old is the patient (in years)? |  |
| What position was the patient in? | • Sitting<br>• Lying<br>• Other/ What was this other position? |
| What was the patient's BMI? | • Low BMI (< 18.5)<br>• Normal BMI (18.5 -29.9)<br>• High BMI (> 30) |
| **Patient Questions** |  |
| What reason(s) did you have for considering using an UniWee? | • NOF<br>• Other type of trauma<br>• Temporary immobilisation post-trauma<br>• Other/ What was this other reason for using an UniWee |
| Have you used an UniWee before this hospital admission? | Y/N |
| Would you recommend using a UniWee to another patient? | • Strongly Agree<br>• Agree<br>• Neither Agree nor Disagree<br>• Disagree<br>• Strongly Disagree |
| What are the advantages of the UniWee? | • reduces pain from movement<br>• privacy (able to manage on my own)<br>• more dignified than a bedpan |
| Was there anything you didn't like about using the UniWee? |  |
| Any other comments |  |
| **Healthcare Staff Questions** |  |
| Have you ever used an UniWee for a patient before | Yes/ No |
| What reason(s) did you have for considering using an UniWee for this patient? | • NOF<br>• Other type of trauma<br>• Temporary immobilisation post-trauma<br>• Other<br>What was this other reason for using an UniWee |
| Did you use an UniWee for this patient? Yes/No | If No: What was the reason(s) for not using an UniWee for this patient?<br>• acute retention/catheter inserted<br>• cognitive status<br>• patient refused |
|  | • I was unsure how to make/use the UniWee<br>• Other/ what was this other reason for not using an UniWee? |
| Would you recommend using the UniWee for your patients | • Strongly Agree<br>• Agree<br>• Neither Agree nor Disagree<br>• Disagree<br>• Strongly Disagree |
| Were there any advantages of using an UniWee? | • Less pain<br>• More dignity<br>• Less work<br>• Avoid catheterisation<br>• Avoided log roll<br>• Avoided soiling<br>• Other/ What were these other advantages?<br>• No, there were no advantages |

#### Study Size

A convenience sample was used. In the final multi-institutional study, each site sought to recruit as many participants as possible with a maximum of 40 patients and 40 healthcare professionals at each site between June 2022 and February 2023 to prevent dominance of any one site in the analysis.

#### Data sources

All data for the multi-institutional study were collected and managed exclusively using Research Electronic Data Capture tools (REDCap) (Harris *et al*. 2009), (Harris *et al*. 2019), a secure, web-based software platform designed to support data capture for research studies. No patient identifiable data was collected. Surveys were considered complete if all questions were answered (except the free text field); incomplete responses were excluded from the analysis database. In some cases where patients were moved out of the Emergency Department before completion of the survey they were contacted retrospectively and consented to participate and complete the survey. (Survey tool available in Supplementary Materials)

Kruskal Wallis test was performed to confirm no significant difference between the age data from the four sites.

Free text comments from both patients and healthcare professionals were thematically analysed using Taguette (Rampin *et al*. 2021). The themes used were negative and positive comments and the negative comment theme included the sub themes of leaks and discomfort.

One way analysis of variance (non-parametric data) of three groups followed by the Friedman test was performed to assess whether patient BMI influenced Healthcare professional participants view of whether they would recommend the urinal.

#### Statistical Methods

Statistical tests were performed using GraphPad Prism version 9.5.1 for Windows, GraphPad Software, San Diego, California USA, www.graphpad.com.

### Reporting Guidelines

SQUIREreporting guidelines were used (Ogrinc et al)

### Ethical considerations

The urinal is a minor modification of a well-established toileting aid that is used in most NHS hospitals. There was no modification to the patients’ surgical or medical management. Verbal consent was sought prior to use and patients without capacity were excluded from the study. Consent was also sought to complete the questionnaire and to use the data in the questionnaire. Data was anonymised prior to processing.

## Results

PDSA cycles (Table 1) allowed us to confirm the utililty and acceptability of a female urinal and to refine the design with reference to comments received. The final cycle involved other institutions to ensure that the urinal was transferable to other hospitals.

Data from the third PDSA cycle are presented below:

*Participants:* A total of 103 patients and 118 healthcare professionals were recruited. *Patient Characteristics:* The age range of the patients was 19 to 102 years (mean= 75, median = 77, n=118). There was no significant difference between the age data from the four sites (p = 0.15). Most patients (81/118) had an estimated normal BMI (18.5-29.9), 12/118 (10%) low BMI (<18.5) and 25/118 (21%) high BMI (>30)

Most patients offered the urinal were immobilised due to a neck of femur fracture 73/118 (62%). Other indications included other lower limb trauma 23/118 (19%), temporary immobilisation post trauma 16/118 (14%) and other issues including personal choice 6/118 (5.0%).

The majority 106/118 (90%) of patients used the urinal in a lying position and 12/118(10%) in a sitting position.

### Patient feedback

#### Utility

The predominate patient reported benefit was that the urinal reduced pain from movement (72/103), followed by it being more dignified than a bed pan (70/103) and increased privacy/ability to autonomously manage (50/103) (Figure 2).

**Figure 2:**
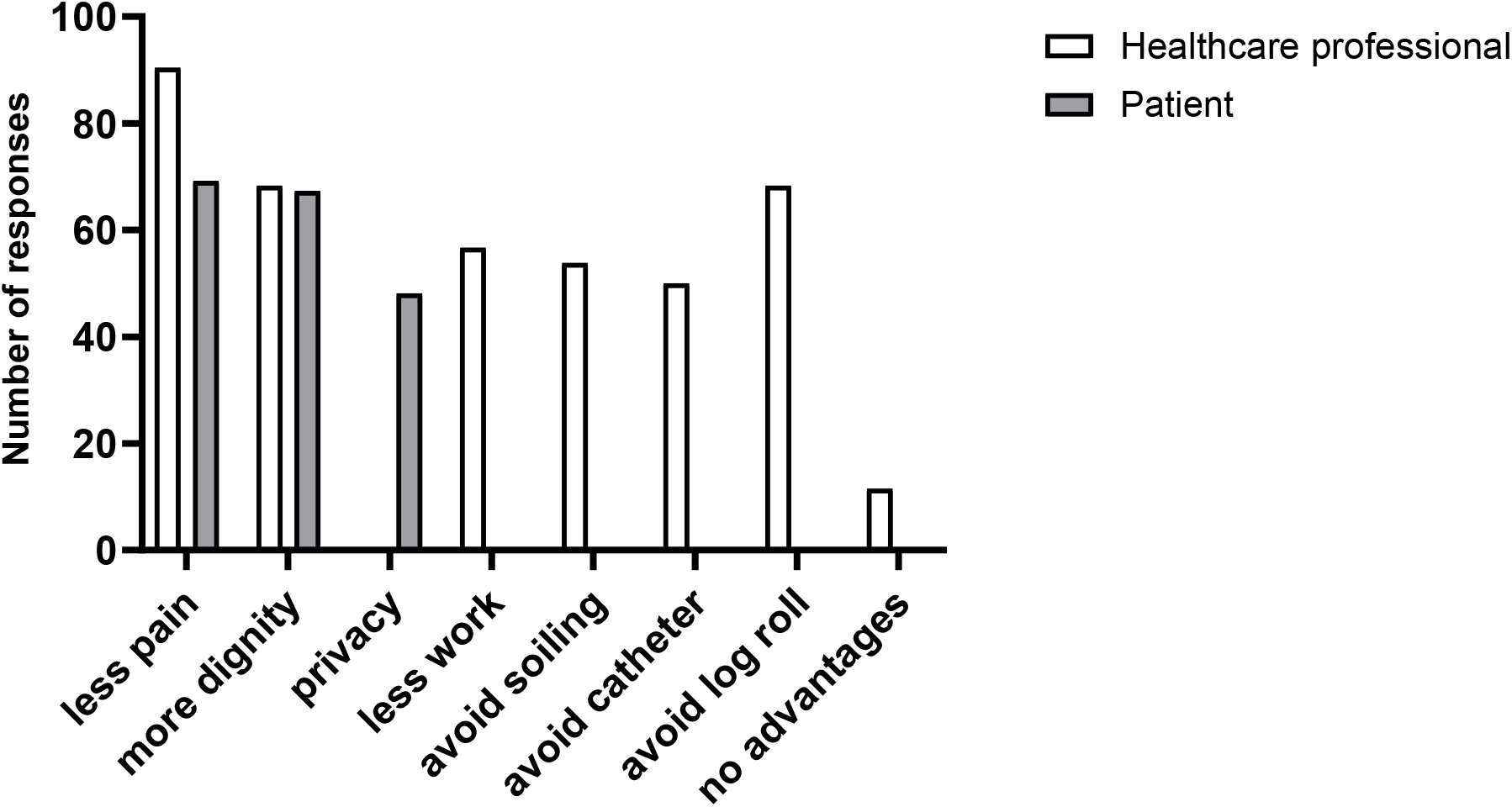
Survey feedback from patients and healthcare professionals (n=118) (N=103) on perceived benefits of using the urinal

#### Acceptability

79% (77/103) of patients strongly agreed (49/103) or agreed (36/103) that they would recommend the urinal to a fellow patient (Figure 3) with 12/103 (11%) strongly disagreeing (6/103) or disagreeing (6/103), with 14/103 neutral responses.

**Figure 3:**
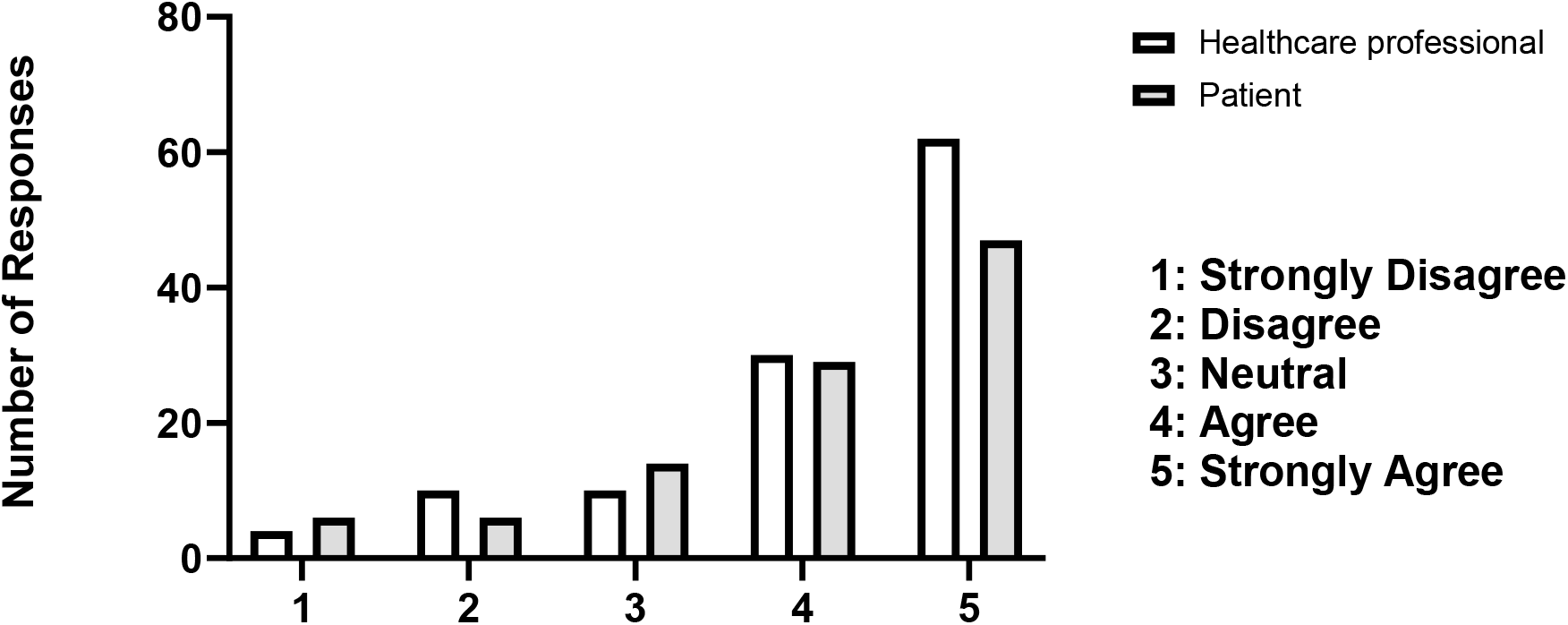
Survey feedback on acceptability of the urinal to patients (n=103) and healthcare professionals (n=118) using Likert scale.

### Healthcare professionals’ feedback

#### Utility

Healthcare professionals perceived benefits (Figure 2) were less pain for the patient (94/118), avoiding the need to log roll a patient (71/118) and increased dignity (71/118). Other benefits included reduction in healthcare professional’s workload (59/118), avoided catheterisation (52/118) and avoided soiling (56/118). 12 responses (12/118) stated that there were no advantages of the urinal over current practice.

#### Acceptability

Most healthcare professionals (94/118) strongly agreed or agreed that they would recommend the use of this urinal to a patient (Figure 3).19% (14/118) strongly disagreed or disagreed and 10/118 were neutral.

### Free text feedback

Fifty-four patients (n=103) provided free text feedback. Thematic analysis of the feedback showed 33 comments were negative and 21 were positive.

- Negative comments included: *‘leakage’, ‘wet’, ‘uncomfortable’*
- Positive comments included: *‘easier / better than bedpan’, ‘brilliant’, ‘should be available in ED’*

31 Healthcare professionals (n=118) provided free text comments of which thematic analysis showed 15 of these were negative and 16 positive.

- Negative comments included: *‘wet’, ‘soiled’, ‘difficult’*
- Positive comments included: *‘less pain’, ‘love this, ‘easy to use’, ‘convenience’*

The data from healthcare professionals’ feedback was analysed to assess whether patient estimated BMI influenced the acceptability of the urinal to healthcare professionals (Figure 4). One way analysis of variance of the three groups followed by the Friedman test showed that BMI significantly (p=0.018) affected the perceived acceptability to healthcare professionals.

**Figure 4:**
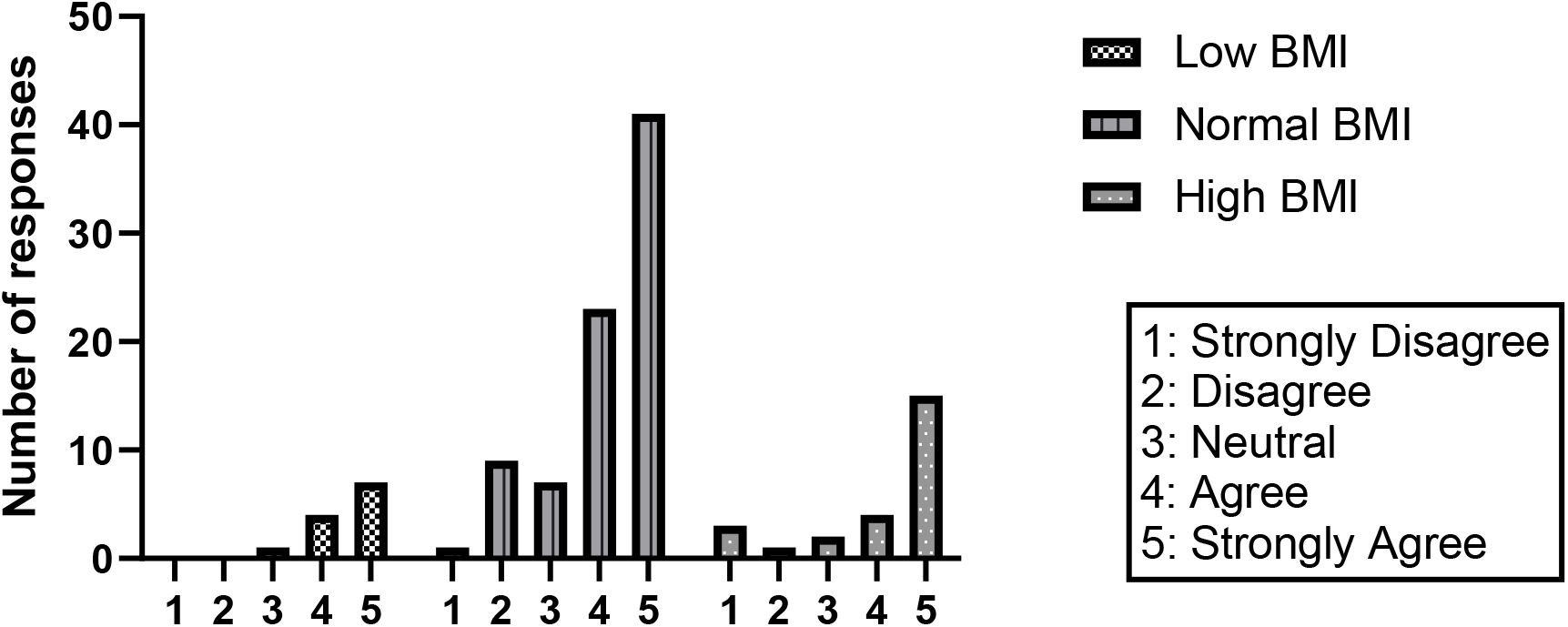
Acceptability of the urinal as a function of estimated BMI (Likert scale).

Initial start-up was dependent on a nursing champion who recruited colleagues to engage in the project. Provision of a design template and clear instructions for creation of the urinal increases uniformity and consistency. There is still inevitable individual variability when making the urinal as it is reliant on tool availability (scissor size and sharpness) as well as staff members skill, interest, and time to ensure smooth edges. Trusts have different suppliers of pulp urinal bottles which vary in terms of design and material composition. The ability to alter the dimensions of the product has potential advantages of customisability in terms of shaping the product to patient habitus, but in turn may lead to the identified issues from free text feedback with cutting, spillage, and rough edges. These can be addressed by the development of a commercially produced, machine-manufactured product.

Sites that had pre-prepared boxes of up to 10-20 products available weekly reported that these were all utilised, demonstrating the actual usage of the urinal was significantly higher than the number of participants recruited.

One site did not routinely use disposable male urinal bottles and did not have an onsite macerator for the disposal of the urine bottles. The use of disposable urine bottles was therefore not imbedded in normal practice and this site recruited least participants.

## Discussion

This project was initiated by a patient using a design developed by our nurses for our patients. Most patients, and healthcare professionals in this trial would recommend the urinal to another patient. Patients felt they suffered less from pain due to movement and that it was a more dignified way to micturate. Healthcare professionals felt that patients suffered less pain related to movement and that there were advantages related to patient urinary care management as the urinal has potential to reduce the need for use of urinary catheters, for lifting and log rolling patients.

There were negative comments relating to leakage and to sharp edges on this handmade device and a lack of confidence in the integrity of the urinal under pressure. Patient estimated BMI had a significant impact on acceptability to healthcare professionals and it was noted that some of the negative comments were related to the urinal not being as effective for patients with a higher BMI.

The NHS Supply chain purchased over 33 million disposable pulp male anatomy urinal bottles in 2022; however 381,624 bottles designed for female anatomy were purchased (personal communication, NHS Supply Chain), despite similar numbers of male and female patients. At the time of the study only one female urinal was available to the NHS, which is a slipper shaped device that operates in a manner similar to that of the bedpan. Another female urinal has recently been added (December 2023) which is a different shape but would be useful as a comparison to a commercial version of this urinal.

The use of this type of urinal addresses an important health inequality, has potential to reduce NHS spend and release staff time. The majority of NHS Trusts in the United Kingdom use disposable pulp products for managing patient waste. The urinal is easily replicable, scalable and can potentially be used in any department in which the pulp urinal bottle and a macerator for disposal are available. We are currently working with a company to make a commercial version of the urinal so that it can be evaluated. It has potential to be used by any patient without male anatomy who is temporarily immobilised; obstetric patients, post operative patients in the recovery, vascular patients pre/post lower limb amputation or patients with poor mobility.

## Supporting information

Participant Survey

## Data Availability

Data are available on reasonable request. Requests to the corresponding author, Siân Thomas (sian.thomas9atnhs.net). De-identified participant data can be made available on request.

## Acknowledgements

We would like to thank the following for their assistance with data collection: Kylie Ashby, Dr Sarah Black, Dr Nell Cuthbertson, Dr Joshua Drinkwater, Jasmin Farooq, James Memory-Ferron, Edel Fritchley, Dr Abdul Hassan, Dr Sherena Jackson, Dr Reem Jaffar, Binta Fatumata Jalloh, Dr Elinor Jones, Ria Osborne, Dr Nahema Rajabali, Dr Shruthi Rayen, Sally Reid-Prescott, Annabel Summers, Dr Kriti Vaidya, Dr Nicola Watson, Dr Benjamin Woolner. We would like to thank the Emergency and Trauma Departments at Gloucestershire Royal Hospital and Great Western Hospital, the Emergency Department at Salisbury District Hospital, the Orthopaedic Department at Torbay Hospital and the South West Ambulance Service for supporting this project enthusiastically without any additional funding.

