## Supplementary material for "A disposable female urinal bottle (the EasyWee ^tm pending^) improves patient experience for immobilised females with lower limb fractures": Participant Survey

### Easywee satisfaction survey

Please complete the survey below.

Thank you!

We have recently started using the Easywee (see picture below), and we want to hear whether or not it is working well for patients and staff. This survey should take 2-3 minutes to complete, and we would love to hear from you

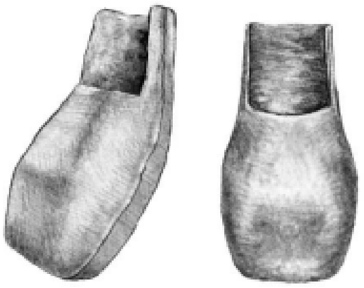

Please let us know that you are happy to complete this survey, and that you are happy for us to use your anonymised feedback

◯ Yes
◯ No

Are you a patient, or a healthcare professional

◯ Patient
◯ Healthcare professional

#### About you and the ambulance service/ hospital

Which ambulance service/ hospital are you a patient in?

- ◯ Gloucestershire Royal Hospital
- ◯ Great Western Hospital
- ◯ Torbay and South Devon Hospital
- ◯ Worcester Royal Hospital
- ◯ Salisbury Hospital
- ◯ SWAST (South West Ambulance Service)

Why did you need to use an Easywee?

- ◯ Hip fracture
- ◯ Other fracture
- ◯ Not allowed to move after trauma
- ◯ During or after giving birth
- ◯ Other

How old are you (in years)?

__________________________________

Which department are you answering from?

- ◯ Trauma and Orthopaedics
- ◯ Obstetrics and Gynaecology
- ◯ Emergency Department
- ◯ Ambulance queue
- ◯ Ambulance

Which ambulance service/ hospital site are you in?

- ◯ Gloucestershire Royal Hospital
- ◯ Great Western Hospital
- ◯ Torbay and South Devon Hospital
- ◯ Worcester Royal Hospital
- ◯ Salisbury Hospital
- ◯ SWAST (South West Ambulance Service)

How old is your patient (in years)?

__________________________________

#### Using the Easywee

| | |
|---|---|
| Have you used an Easywee before this hospital admission? | ◯ Yes<br>◯ No |
| Would you recommend using a Easywee to another patient? | ◯ Strongly Agree<br>◯ Agree<br>◯ Neither Agree nor Disagree<br>◯ Disagree<br>◯ Strongly Disagree |
| What are the advantages of the Easywee?<br><br>(please select all that apply) | ☐ reduces pain from movement<br>☐ privacy (able to manage on my own)<br>☐ more dignified than a bedpan |
| Was there anything you didn't like about using the Easywee? | ________________________________ |
| Any other comments | ________________________________________ |
| Have you ever used an Easywee for a patient before | ◯ Yes<br>◯ No |
| What reason(s) did you have for considering using an Easywee for this patient?<br><br>(please select the best answer) | ◯ NOF<br>◯ Other type of trauma<br>◯ Temporary immobilisation post-trauma<br>◯ Pre/postnatal care<br>◯ Other |
| What was this other reason using an Easywee | ________________________________ |
| Did you use an Easywee for this patient? | ◯ Yes<br>◯ No |
| What was the reason(s) for not using an Easywee for this patient?<br><br>(please select all that apply) | ☐ acute retention/catheter inserted<br>☐ cognitive status<br>☐ patient refused<br>☐ I was unsure how to make/use the Easywee<br>☐ Other |
| What was this other reason for not using an Easywee? | ________________________________ |

#### About your patient

| | |
|---|---|
| Did your patient provide feedback? | ◯ Yes<br>◯ No |
| Reason: (e.g. left department ) | __________________________________<br>(as per comp_unable) |
| What position was the patient in? | ◯ Sitting<br>◯ Lying<br>◯ Other |
| What was this other position? | __________________________________ |
| What was the patient's BMI?<br><br>(an estimate is fine) | ◯ Low BMI (< 18.5)<br>◯ Normal BMI (18.5 -29.9)<br>◯ High BMI (> 30) |
| Would you recommend using the Easywee for your patients | ◯ Strongly Agree<br>◯ Agree<br>◯ Neither Agree nor Disagree<br>◯ Disagree<br>◯ Strongly Disagree |
| Were there any advantages of using an Easywee?<br><br>(please select all that apply) | ☐ Less pain<br>☐ More dignity<br>☐ Less work<br>☐ Avoid catheterisation<br>☐ Avoided log roll<br>☐ Avoided soiling<br>☐ Other<br>☐ No, there were no advantages |
| What were these other advantages? | __________________________________________ |
